# Near Real-Time Measures of Interregional Mobility Restrictions on COVID-19 Transmission

**DOI:** 10.1101/2024.06.14.24308359

**Authors:** Jesús Berríos, César Marín Flores, Bernardo Gutierrez, Loreto Bravo, Leo Ferres

## Abstract

The global SARS-CoV-2 pandemic prompted nations to implement mobility limitations to curb virus spread. In Chile, targeted interregional measures were employed to mitigate the social and economic costs. Here, we employ a novel real-time methodology to assess the impact of such mobility restrictions on epidemic control. Leveraging telecom-derived eXtended Detail Records (XDR) and official COVID-19 epidemiological data, we estimate interregional mobility and disease prevalence. Employing Bayesian adjustments, we compare different mobility restriction scenarios: business-as-usual (BAU), initial measures, and total lockdown. Mobility reductions significantly curtailed cases and risk across regions. Even modest mobility declines under total lockdowns considerably lowered imported cases. The high-risk Santiago Region, a national source of infections to other regions, demonstrated lowered risk due to mobility restrictions. Our approach facilitates rapid regional insights for informed policy responses.

**Author summary:** The implementation of mobility restrictions is a coarse approach to limiting pathogen transmission during the early stages of an epidemic. Disentangling the effects of the reduced population mixing and the effects of new seeding events can be challenging to perform in real time, making any evaluation of the efficacy of such interventions a post hoc exercise. Here we present an approach that uses aggregated data from individual mobile phone locations within the national network of antennas to statistically correct for changes in human mobility and estimate the changes in COVID-19 transmission across regions in Chile during the first wave of the pandemic. We estimate a risk score that can be implemented nearly in real time to estimate the effects of mobility restrictions on viral transmission and highlight that important urban centres that seed other regions, like the Santiago Region, are at low risk of experiencing increased viral transmission due to new seeding events from other regions.

## Introduction

The initial genetic sequencing of the SARS-CoV-2 virus on January 12, 2020, marked the beginning of a rich body of multidisciplinary research [1]. Initial inquiries focused on viral propagation characteristics and under-reporting rates [2, 3, 4, 5]. This was soon followed by studies that explored the effects of restricting international travel and domestic mobility [6, 7, 8, 9].

Efforts to pinpoint inaccuracies in real-time epidemiological assessments have identified several challenges. These include the prevalence of asymptomatic cases [2, 10, 11], variability in incubation periods [12], and increased transmissibility during early infection [13]. Consequently, policymakers have had to evaluate the costs and benefits of various interventions, such as case isolation and contact tracing, the effectiveness of which has been variable [14].

Bayesian models have been developed to address these complexities [3, 4] which use more robust indicators, like death counts, to estimate the pandemic’s reach. In the Chilean context, the model in focus [4] assesses daily variations in the Case Fatality Ratio (CFR), adjusted by the hospitalization-to-death interval [5]. This approach has shown consistent results across countries [4, 7].

Demographic adjustments to the base CFR are crucial for accurate modeling due to age-related variance in case severity [15]. This aspect is particularly relevant in Chile, where the age distribution of the population can vary significantly across its regions.

Chile’s strategy against SARS-CoV-2 heavily relies on non-pharmaceutical interventions (NPIs) like social distancing and quarantines, influencing domestic mobility patterns [16, 17]. Studies affirm that travel restrictions, while effective in controlling viral spread [8, 9], have significant economic ramifications [18]. Their success is contingent upon various factors such as timing, duration, compliance, and local conditions [9, 4].

Mobile phone data have been instrumental in evaluating mobility trends in Chile, serving multiple applications from transportation analytics [19] to social inequality studies in the context of COVID-19 [18]. These data have enabled precise tracking of NPI impacts at community, regional, and national levels.

Building on this existing research, our study aims to extend these insights to broader geographical settings and conditions within Chile. We focus on interregional movement, the efficacy of restrictions, and viral dynamics, contributing to the global knowledge base on managing the ongoing SARS-CoV-2 pandemic. Our study introduces a robust methodology for unravelling regional COVID-19 transmission dynamics and informing real-time policy in a context of diverse demographics and centralized resources. Santiago Region serves as an illustrative case of a SARS-CoV-2 net exportation hub, accentuating the importance of risk ratings of new viral importations as a key metric in evaluating transmission related to mobility. Our analysis unveils that maintaining pre-pandemic mobility would necessitate stricter interventions due to elevated risk ratings, elucidating the efficacy of NPIs at a granular, regional level. The complexities of transmission dynamics, as highlighted by the Metropolitan region’s centrality in nationwide viral dissemination, reinforce the inadequacy of a blanket approach to public health interventions. These insights collectively offer a data-driven framework for navigating the intricacies of pandemic management, setting the stage for future investigations that aim to integrate additional variables such as vaccination rates or healthcare infrastructure.

## Methods

### Data Acquisition and Processing

This study relies on three primary data sources: mobile telecommunications records (XDRs), the 2017 Chilean population census, and the official COVID-19 statistics from the Chilean government. We sourced mobile data from Telefónica/Movistar Chile, which accounts for 29-32% of Chile’s mobile subscriber market. The eXtended Detail Records (XDR) log traffic transactions for all users. Each XDR is represented as a tuple (*u, t, A*), where *u* is an anonymized user identifier, *t* is the transaction timestamp, and *A* is the associated antenna. These antennas, with fixed geographical locations, track user movements across the 16 official regions of Chile which are the country’s first-level administrative division. For each date *d* from March 1, 2020, to August 31, 2020, and for every regional pair *r*_1_ and *r*_2_, we counted devices *n* based on their initial detection in *r*_1_ and subsequent presence in *r*_2_. Further details regarding the mobility dataset can be found in [20]. Population data was sourced from the 2017 Chilean Census [21]. We also used expansion factors for our analysis [22]. In the context of inter-regional travel, regional population distribution is imperative for determining the prevalence of origin regions. The Ministry of Science, Technology, Knowledge, and Innovation updates the national COVID-19 metrics. We utilized the datasets ‘Total New Cases by Region’ and ‘COVID-19 deaths by Region’ [23].

### Methodology

In the characterization of interregional mobility, especially considering the Non-Pharmaceutical Interventions (NPIs) that introduced mobility constraints, we define three distinct study scenarios. The **Baseline Mobility (BAU - Business as Usual)** represents interregional origin-destination travel data spanning a 10-day interval before the advent of the pandemic, specifically from March 1 to March 10. The **Initial Measures Scenario** captures a subsequent 10-day period, from May 8 to May 17, showcasing reduced travel compared to the baseline, marking the period when the first NPIs were instituted. Lastly, the **Complete Quarantine Scenario** focuses on data from May 23 to June 1, a phase during which a comprehensive quarantine was enforced in the Metropolitan Region, Chile’s most densely populated region [18].

#### Calculation of the Corrected Case Fatality Ratio

Our analysis starts with the estimation of the baseline Case Fatality Ratio (CFR), which is expected to be compared with the actual numbers of cases, across various Chilean regions. In contrast to the approach taken by Russell et al. [4], our methodology incorporates age-specific CFR estimates as described by Verity et al. [15], and adapts them to the unique demographic composition of each region as suggested by [24]. Specifically, our model applies cumulative age-related fatalities, derived from a calibrated age-dependent CFR. This ratio is set in proportion to the maximum original CFR found in the oldest age group. The model’s performance is then assessed against observed age-related death patterns through a Poisson likelihood analysis. Subsequently, the corrected daily CFR is calculated using a delay distribution, which adjusts the number of daily reported cases in the regions to their known outcomes (*dC*_*r,t*_) for a subsequent time *t*, through a method of discrete convolution correction. The chosen distribution corresponds to Hospitalization to Death [5], modeled as a lognormal distribution with a mean of 13 days (95% CI 8.7 - 20.9 days) and a standard deviation of 12.7 days. Then, for each day, the ratio of the incidence of deaths to the resultant value of the convolution is computed to yield the new corrected CFR (*cCFR*_*c,t*_). The ratio between the stratified base CFR and the corrected cCFR was computed and used as an estimate of COVID-19 case detection assertiveness in different Chilean regions. This primary indicator provides insight into the proportion of symptomatic cases effectively identified by health authorities. It is important to note that the results of case assertiveness calculation at the regional level are constrained to regions that have accumulated at least 10 COVID-19 deaths; this constraint was adopted from the methodology in [7]. The point calculation of case assertiveness lacks robustness for assertiveness rates that vary over time. As a result, a Gaussian Process described in [4] was implemented, taking into account these initial estimations of Case Fatality Ratio for the likelihood function of the model.

#### Calculation of Incidence, Prevalence, Imported Cases, and Risk Rating of new viral importations

First, infections are estimated from the case assertiveness curve for each region. This involves relating the number of daily reported cases to the case assertiveness index per day and then dividing by the proportion of asymptomatic cases, which was considered at 50% with a confidence interval [10%, 70%], the same proportion and interval used in the study [7]. To derive the incidence estimate for each defined destination region, the average daily infections within a specified time interval are computed.

The prevalence of cases in the origin regions was estimated on a daily basis. Initially, the rolling mean of the adjusted cases by assertiveness indices and asymptomatic rates over the previous 10 days is calculated for each day, and subsequently averaged and converted to a proportion by dividing by the regional population, which will result on a prevalence estimate for each region in each scenario of study. Moving on to the calculation of case imports, a key assumption is made to adjust the trips by the market share of mobile subscribers, as the travel sample represents 29-32% of mobile subscribers in Chile according to [6]. Thus, this proportion was approximated to 0.3, and the total number of trips was divided by this value to estimate the overall number of trips. Case imports were thereafter estimated by multiplying the prevalence value by the adjusted number of trips for each destination region and averaging the resulting values within each scenario’s time window for each region.

Next, the risk rating of new viral importations was calculated by dividing the average expected imported cases by the average of the adjusted daily cases, which represents the local incidence for each region during the study period. The results were stratified into three risk ranges: less than 1%, between 1% and 10%, and greater than 10%. These values determine the effect of imported cases due to interregional travel for each region.

Finally, the Basic Reproductive Number (*R*_*t*_) for each region was estimated using the publicly accessible EpiEstim tool, available as an R package. This resulting Rt does not consider potential imported cases estimated in previous steps. EpiEstim allows for quantifying the transmissibility of a pandemic based on incidence curves as described in [24]. The estimation process was conducted parametrically, using case incidence data obtained from the official sources of the Ministry of Health and a serial interval described by [23] as a normal distribution with a mean of 5.29 days and a standard deviation of 5.32 days.

## Results

At the crux of the investigation into the dynamics of interregional mobility restrictions on COVID-19 transmission in Chile lies the nuanced assertiveness indices. Figure 1 shows these indices calculated for different regions and illustrates the fluctuations in confirmed cases and mortality statistics concerning a baseline Case Fatality Rate (CFR) value. The approach outlined in [6] utilizes a unique CFR value for Chile of 1.4 [1.2, 1.7]; however, it is imperative to acknowledge that the CFR baseline may undergo variations owing to the distinct demographic characteristics within each regional population. Our approach which stratifies these CFR values according to age and population improves on the precision of these estimates. The onset of assertiveness indices is not uniform across regions due to these being constrained to instances where at least 10 confirmed COVID-19 deaths have been reported. As a result, assertiveness indices for regions like Arica and Parinacota or Los Ríos are not available until after June, while others like Aysén and Atacama are entirely unavailable due to insufficient data. These indices are sensitive to abrupt changes in official reports, as they are derived from a ratio of baseline and adjusted CFR values—falling below 1 in regions like Tarapacá and Antofagasta and exceeding 1.5 in Valparaíso and Ñuble. The country exhibited fluctuating assertiveness in the initial months of the pandemic. The performance of the Metropolitan Region aligns closely with the national total (the panel labeled “Chile” in Figure 1) during this time frame. This alignment is likely attributable not only to the region’s high population concentration but also to its earlier epidemic onset compared to other areas, particularly in the northern part of the country.

**Figure 1:**
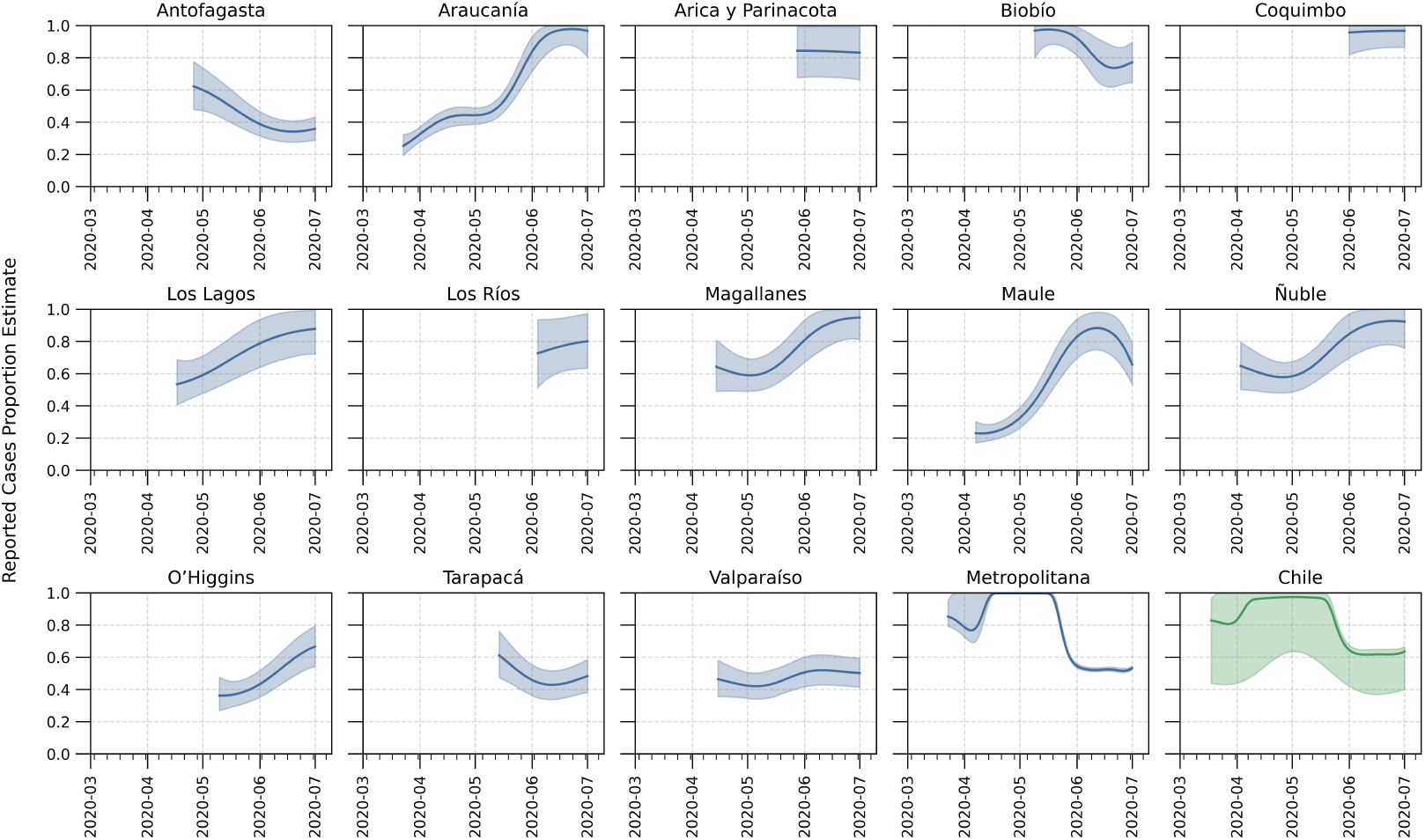
Temporal Evolution of Case Assertiveness Indices Across Chilean Regions: Estimated time-varying proportion of reported COVID-19 cases as a function of date are shown, produced using a Gaussian process model. Each panel represents a different region of Chile, illustrating how the assertiveness index varies over time. The solid blue line is the estimated median proportion of symptomatic cases ascertained over time and the shaded blue region is the 95% credible interval of these ascertainment estimates. The assertiveness index serves as a measure of the effectiveness of case reporting and detection within each region. The last subfigure, in green, shows the results for all of Chile.

Interregional travel patterns, depicted in Figure 2, reveal higher mobility rates in the scenario preceding non-pharmaceutical interventions (NPIs). A visual examination confirms reduced mobility between the Business-As-Usual (BAU) scenario and initial measures (partial lockdown). However, the reduction is less pronounced between the latter and the total quarantine (full lockdown) in Santiago and other regions.

**Figure 2:**
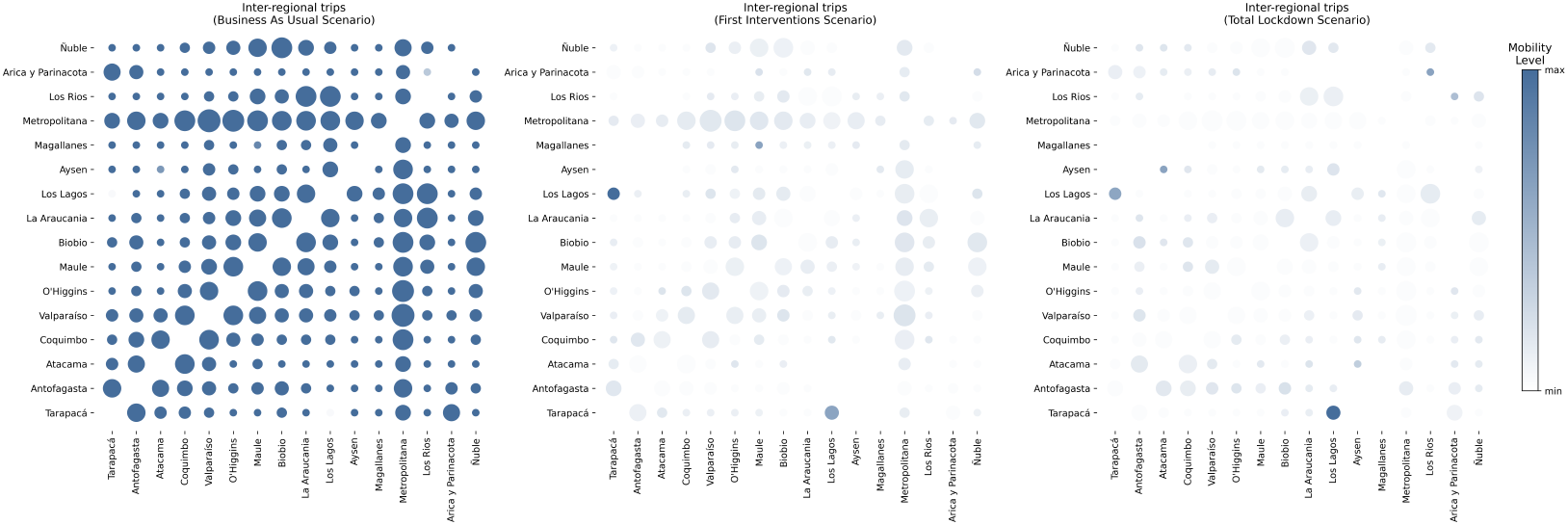
Interregional Mobility Matrix Across Three Examined Scenarios: Inter-regional mobility patterns during three different mobility scenarios are shown. Circle diameters indicate the number of trips between regions, whereas the opacity indicates the level of mobility regarding the other scenarios, with darker shades signifying higher levels of mobility. Panel A represents the Business-As-Usual (BAU) scenario, where no restrictions are in place. Panel B depicts the situation under partial lockdown, reflecting the effects of initial restrictive measures. Panel C shows the conditions during a total lockdown, illustrating the most severe restrictions on mobility.

The visualization in Figure 2 illustrates interregional mobility patterns, modulated by opacity levels. An outlier in this dataset can be detected, requiring further investigation, is the behavior between Los Lagos and Tarapacá, where travel increased by three orders of magnitude under restriction scenarios. However, this anomaly does not impact subsequent analyses and results.

Travel to and from the Metropolitan Region significantly diminishes, nearly in its entirety, in the third scenario. This is consistent with the expected effects of the lockdown implemented in the region after May 17, 2020. At this critical time, the region had a high number of confirmed cases and assertiveness gradually fell from 100% to 50%. While a clear geographic pattern is observed, where proximal regions exhibit high mobility between them and reduced mobility to distant regions, the Metropolitan Region is an exception: despite its distance from Aysén, Los Lagos, and Antofagasta, the region exhibits a high total number of trips2.

The evaluation of risk ratings of new viral importations per region across multiple scenarios can play an instrumental role as a quantifiable measure of the ratio of imported cases to local incidence. Upon comparing the Initial Measures (I) and Total Lockdown (II) scenarios (Figure 3, left map for each box), it becomes evident that risk ratings increase across regions. In the first scenario, 5 out of 11 regions had a risk rating below 1%; this number decreased to 3 out of 13 regions in the second scenario, warranting more stringent interregional mobility restrictions. This assertion is corroborated by the fact that 77% of regions with available data in the second scenario exhibited a risk rating exceeding 1%, significantly impacting local incidence rates for a substantial portion of the population.

**Figure 3:**
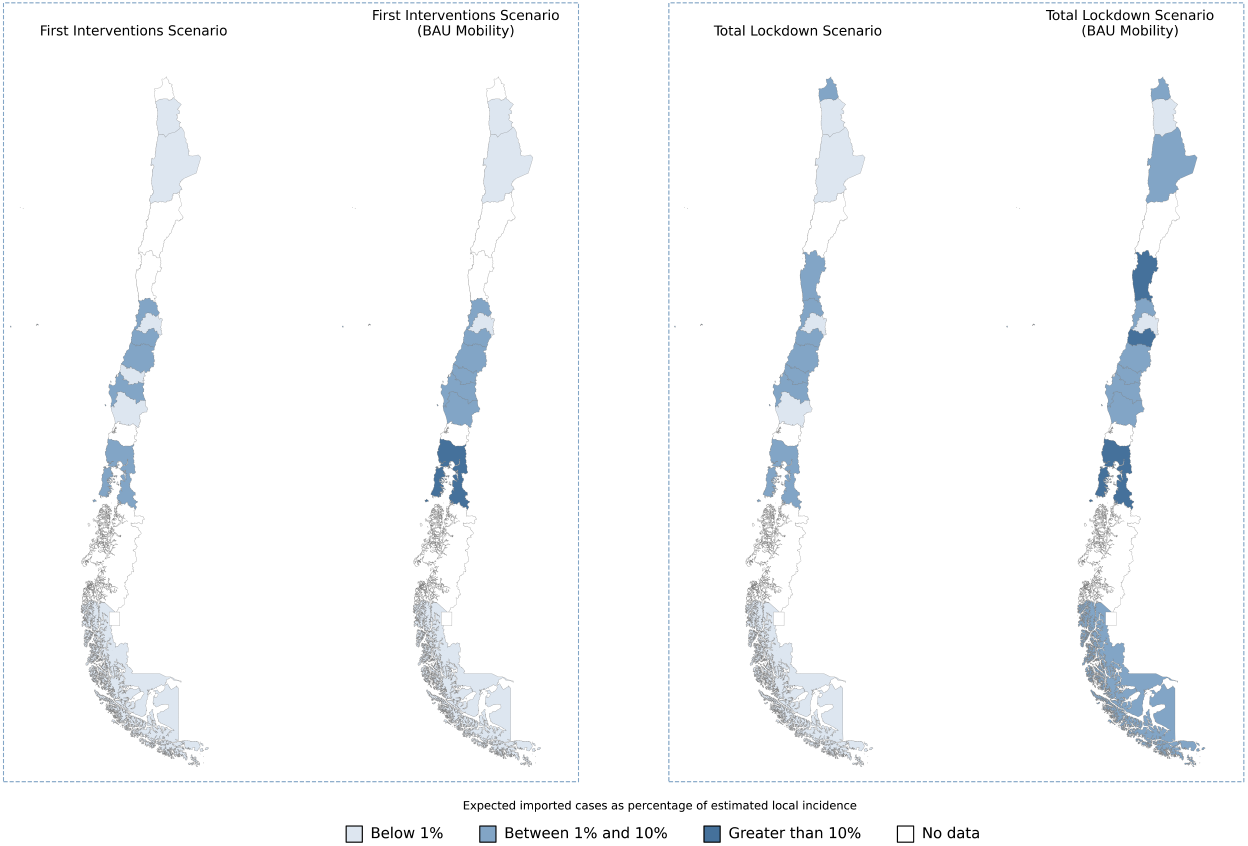
Regional Risk Ratings of new viral importations in the First Interventions and Total Lockdown Scenarios, considering the mobility for each scenario and Business-As-Usual (BAU): Risk ratings are categorized into three segments —0-1%, 1-10%, and greater than 10%— to facilitate a visual elucidation of the impact of imported cases on local incidence rates across a spectrum of mobility and non-pharmaceutical interventions scenarios.

Contrasting these scenarios with a pre-pandemic Business-As-Usual (BAU) mobility model reveals that interregional travel restrictions are further justified. Not only do multiple regions exceed a 1% risk rating for case importations, but several also reach values exceeding 10%. This assessment underscores the need for stringent measures. Notably, the Metropolitan Region, despite a high local incidence of COVID-19, can maintain this risk rating consistently below 1%, except for scenario I with BAU mobility. This indicates that although the Metropolitan Region could be impacted by imported cases, the local incidence is sufficiently high to mitigate increased risk from such cases.

The assessment of COVID-19 cases exported from each region to the broader national landscape explores the repercussions of interregional mobility on the composition of imported cases. This analysis (Figure 4), shows that the Metropolitan Region plays a crucial role in exporting cases nationally, specifically within the context of the First Interventions and Total Lockdown scenarios, contributing a significant proportion of imported cases to other locations. Specifically, in regions such as Valparaíso, Biobío, O’higgins, Los Lagos, and Coquimbo—each already having a high case count—the Metropolitan Region accounted for at least 85% of imported cases explained by the high prevalence and mobility from this region during both periods.

**Figure 4:**
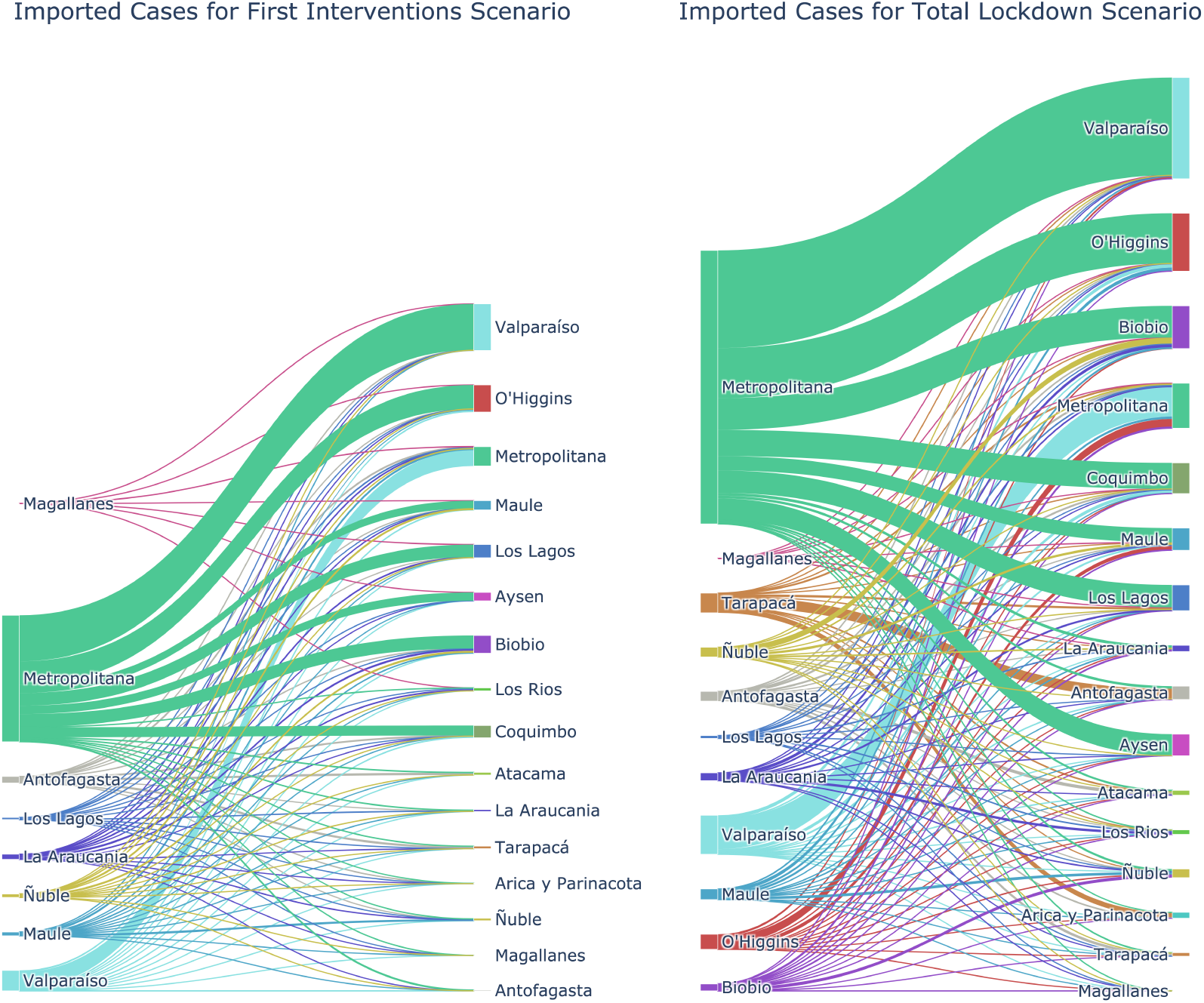
COVID-19 Regional Case Importation Patterns in the First Interventions and Total Lockdown Scenarios. The Sankey diagram presents the intricate dynamics of case importations between geographical regions, comparatively illustrating the viral spread across regions through importation patterns between the First Interventions and Total Lockdown scenarios.

Figure 4 demonstrates that the Metropolitan Region is a key driver in exporting COVID-19 cases to the rest of the country. In regions like Valparaíso, Biobío, O’Higgins, Los Lagos, and Coquimbo, which already had a significant number of total cases, the Metropolitan Region accounted for the majority of imported cases. Comparing case exportation between scenarios reveals that the distribution of positive case flow remains largely consistent, with the Metropolitan Region being the main contributor to the spread of the virus across Chile. Moreover, a general increase in the flow of imported cases among different regions is observed. It’s noteworthy that this increase is not attributable to a rise in interregional mobility, as Figure 2 shows a reduction in mobility between the Initial Measures and Total Quarantine scenarios. Therefore, the uptick in imported cases can be attributed to the growth in the number of cases in all regions, which directly impacts the prevalence value in the originating regions.

Within the established methodological framework in [7], the derived risk ratings of new viral importations and *R*_*t*_ estimates collectively delineate the essential criteria for formulating interregional mobility restriction policies. Figure 5 presents the results of the actual situation of the first interventions and the estimations under BAU mobility. The general trend when comparing both epidemiological scenarios is an increase in the risk rating in all regions under BAU mobility. Additionally, it is observed that only in 2 out of the 11 studied localities, interregional mobility restriction measures would not have an impact on epidemic control.

**Figure 5:**
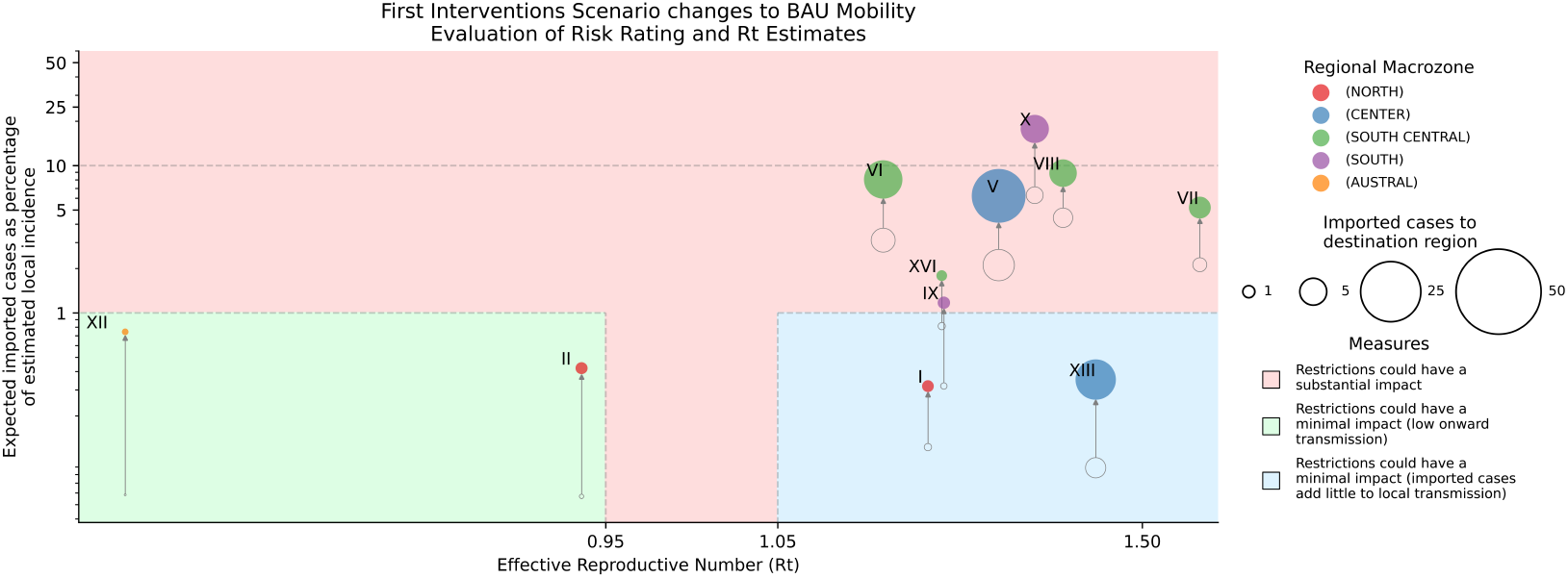
Pandemic situation shifts from the actual first interventions scenario to the estimations under BAU mobility. In accordance with the methodology outlined in [7], mobility restrictions are evaluated as follows: minimal impact when Rt is below 0.95 with a risk rating under 1%, and when Rt exceeds 1.05 with a risk rating under 1%, as imported cases insignificantly affect local incidence in these scenarios. Substantial restrictions are warranted if the risk rating surpasses 1% or, for risk ratings below 1%, Rt falls within [0.95, 1.05], signifying a potential turning point in the epidemic where external incidences could significantly influence regional pandemic situations.

**Figure 6:**
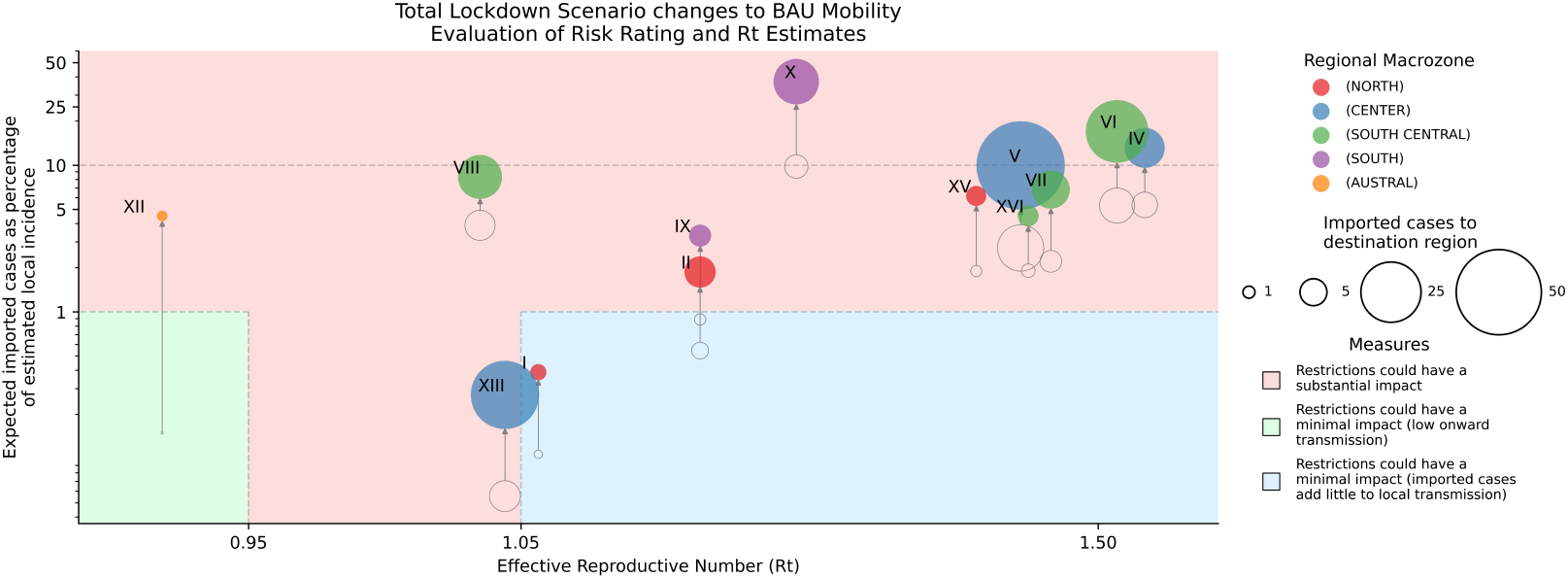
Pandemic situation shifts from the actual total lockdown scenario to the estimations under BAU mobility. In accordance with the methodology outlined in [7], mobility restrictions are evaluated as follows: minimal impact when Rt is below 0.95 with a risk rating under 1%, and when Rt exceeds 1.05 with a risk rating under 1%, as imported cases insignificantly affect local incidence in these scenarios. Substantial restrictions are warranted if the risk rating surpasses 1% or, for risk ratings below 1%, Rt falls within [0.95, 1.05], signifying a potential turning point in the epidemic where external incidences could significantly influence regional pandemic situations.

The entire Central-South macrozone consistently exhibits risk indicators surpassing the critical 1% threshold for both scenarios, with O’Higgins (VI) and Biobío (VIII) notably soaring beyond 5% for BAU mobility estimations. This starkly underscores the imperative for stringent restrictions within this geographical expanse, given its profound impact on pandemic control. Valparaíso (V Region) emerges as the epicenter for total imported cases, resulting in a risk rating exceeding 5% for BAU mobility. According to the methodology [7], this situation strongly advocates for implementing interregional mobility restrictions to curtail its considerable influence on the progression of the epidemic in the country. In contrast, its adjacent region, the Metropolitan Region (XIII), witnesses a rising transmission rate (*R*_*t*_ *>* 1.05) in both scenarios, which diminishes the potential for imported cases to significantly contribute to local spread.

The alterations in risk ratings and the influx of cases from the real total lockdown scenario to the projections under the BAU mobility scenario are portrayed in 6, where the number of daily imported cases increases in contrast to Figure 5 which examines the first intervention scenario. Furthermore, it becomes apparent that no region falls within the “green zone”, where the impact of mobility restriction measures is minimal.

The Metropolitan Region (XIII) along with Tarapacá (I) are the only ones that would maintain a risk rating below 1% under a BAU mobility scenario. The rest remain above 5%, with even four regions surpassing 10%.

Furthermore, it should be noted that the epidemiological scenario during the lockdown had become more complex when compared to the one studied during the initial mobility interventions: only the Magallanes Region (XII) starts within the green zone under the real epidemiological and mobility scenario. However, under the BAU estimates, this changes to a scenario where intervention measures have a substantial impact.

## Discussion

Our study introduces a comprehensive methodology that provides an understanding of regional virus transmission dynamics and informs near real-time policy decisions. This is particularly significant in the context of a country characterized by diverse demographics and a concentration of resources and population in its capital city. To that end, the Santiago (Metropolitan) Region emerged as a critical locus of COVID-19 case exportation, providing an instructive case study that informs our broader understanding of interregional virus transmission.

Risk ratings proved to be indispensable in our analysis, serving as a robust metric for evaluating the dynamics of COVID-19 transmission vis-à-vis interregional mobility. These ratings allow us to quantify the ratio of imported cases to local incidence, which has the potential to significantly impact regional risk assessments. These ratios are particularly important during early stages of an epidemic when transmission dynamics can be mostly influenced by new introductions. The policy implications of such an approach are noteworthy; they provide health officials and policymakers with valuable insights to guide the design of interregional mobility restrictions. These are crucial for controlling viral spread during an epidemic’s early stages under a containment strategy, and thus better managing the epidemic in the country.

In evaluating the effectiveness of Non-Pharmaceutical Interventions (NPIs), our study further indicated that maintaining pre-pandemic levels of mobility would escalate the risk ratings across regions. Such a scenario would necessitate the imposition of stringent measures, entailing far-reaching socio-economic consequences. Here, the integration of mobile phone records and official COVID-19 data proved invaluable, enabling a nuanced assessment of NPI efficacy at a regional and decentralized scale.

The study also highlighted the complexity involved in assessing the influence of restrictions on transmission dynamics. These dynamics can be impacted by a variety of factors such as the rate of onward transmission and the extent to which imported cases contribute to local spread. Our analysis underscores the necessity of considering these complexities when formulating public health strategies.

Notably, the Metropolitan region emerged as a significant contributor to the nation-wide pool of exported COVID-19 cases. This region’s unique demographic and geographic attributes make it a pivotal player in the spread of the virus. The region’s high population density and role as an economic and transport hub substantiates its centrality in the network of viral transmission. This finding further emphasizes the significance of strategic planning in interregional mobility and public health interventions.

In conclusion, our study delineates a multi-faceted landscape of COVID-19 transmission dynamics, bridging the gap between data analytics and actionable policy insights. The rich tapestry of factors affecting virus spread, from regional characteristics to the effectiveness of NPIs, underscores the value of an integrated, data-driven approach in navigating the complexities of pandemic management. Future work should aim to extend this framework to incorporate additional variables, such as vaccination rates or healthcare infrastructure, to create an even more robust decision-making tool.

## Data Availability

Data and code related to the analysis and figures in tis manuscript are publicly available at GitHub. Raw mobility data from individual mobile phones cannot be made publicly available to protect the individual privacy of users.

https://github.com/jesusfberrios/covid_rt_reg_restr.git

## Data and code availability

Data and code related to the analysis and figures in this manuscript are publicly available at our GitHub public repository. Raw mobility data from individual mobile phones cannot be made publicly available to protect the individual privacy of users.

## Ethics statement

This study utilises anonymized, unidentifiable data collected from individual mobile phone devices and publicly available aggregated case count data in Chile. It does not use or access individual personal information, and has therefore been waived for an ethical approval. This decision was made by the Institutional Ethics Committee at Universidad del Desarrollo, Santiago de Chile, Chile, communicated to the authors on March 30, 2022.

## Acknoweledgments

LF and LB thank the funding and support of Telefónica R&D Chile and CISCO Chile. This research was supported by FONDECYT Grant N°1130902 to Loreto Bravo and FONDECYT Grant N°1221315 to Leo Ferres. BG acknowledges funding from the Oxford Martin School Pandemic Genomics programme and the European Union Horizon 2020 MOOD (#874850). LF also acknowledges financial support from the Lagrange Project of the Institute for Scientific Interchange Foundation (ISI Foundation), funded by Fondazione Cassa di Risparmio di Torino (Fondazione CRT).

## Author contributions

LF and LB conceptualized the work. LF, JB and CMF performed data mining and curation. LF, JB and CMF carried out the formal analyses. JB, and CMF produced visualizations. JB, CMF, LF, LB and BG prepared the manuscript. All authors edited and and reviewed the final version.

